# C-Reactive Protein-Triglyceride-Glucose Index, Inflammatory-Metabolic Burden, and Prevalent Stroke in U.S. Adults: NHANES 1999-2010

**DOI:** 10.64898/2026.09.07.26362446

**Authors:** Qile Deng, Zhao Yin

## Abstract

**Background:** The C-reactive protein-triglyceride-glucose index (CTI) integrates inflammatory and metabolic markers, but recent studies have already linked CTI to stroke and cardiovascular outcomes. Whether CTI provides information beyond established risk factors in the U.S. general population remains uncertain.

**Methods:** We analyzed 11,809 adults from NHANES 1999-2010 with fasting laboratory, stroke, covariate, and survey-design data. Stroke was defined by self-reported physician diagnosis. Survey-weighted logistic regression estimated odds ratios (ORs) and 95% confidence intervals (CIs). We evaluated restricted cubic splines, exploratory subgroup interactions, and sensitivity to excluding CRP >10 mg/L. CTI was compared with TyG and its component biomarkers, and incremental value was quantified using weighted C-statistics, integrated discrimination improvement, and continuous net reclassification improvement.

**Results:** Among 11,809 participants, 411 reported stroke. CTI was associated with prevalent stroke before cardiometabolic adjustment (unadjusted OR = 2.05, 95% CI: 1.74-2.42; demographically adjusted OR = 1.57, 95% CI: 1.31-1.89; both P < 0.001), but the fully adjusted association was attenuated (OR = 1.22, 95% CI: 0.98-1.53; P = 0.080). There was no evidence of non-linearity, and subgroup interaction tests, including the CTI-by-age interaction, were not statistically significant. Adding CTI to traditional covariates minimally changed discrimination (weighted C-statistic, 0.846 to 0.847).

**Conclusions:** CTI reflects an inflammatory-metabolic burden phenotype, but its association with prevalent stroke was largely accounted for by traditional risk factors, and it provided negligible incremental discrimination in this cross-sectional analysis.

## Introduction

Stroke is a major cause of death and long-term disability, and its population burden rises with age, obesity, hypertension, diabetes, and related cardiometabolic conditions[1, 2]. Stroke reflects accumulated cerebrovascular injury involving endothelial dysfunction, arterial remodeling, and metabolic stress across the life course[3]. Inexpensive and routinely available biomarkers may help characterize the inflammatory-metabolic burden associated with prevalent stroke at the population level.

Chronic low-grade inflammation and metabolic dysregulation contribute to vascular vulnerability. Inflammation, insulin resistance, triglyceride-rich dyslipidemia, and impaired glucose homeostasis contribute to atherosclerosis, endothelial dysfunction, thrombotic activation, and cerebrovascular injury[4, 5]. C-reactive protein (CRP) is a widely used marker of systemic inflammation and has been evaluated in relation to vascular outcomes[6–8]. Triglycerides and fasting glucose capture lipid and glucose metabolism, and the triglyceride-glucose index has been used as a practical surrogate of insulin resistance and cardiometabolic burden[9]. A composite marker that incorporates these domains may summarize correlated inflammatory-metabolic burden.

The C-reactive protein-triglyceride-glucose index (CTI) combines CRP, triglycerides, and fasting glucose and was originally developed as a cancer-survival prognostic index[10]. Subsequent studies have associated CTI with coronary heart disease, cancer-related outcomes, depressive symptoms, erectile dysfunction, and cardiovascular outcomes[11–18]. More recent longitudinal analyses have examined baseline CTI, cumulative exposure, or trajectories in relation to stroke and cardiovascular events in Chinese cohorts, and NHANES studies have assessed CTI in selected high-risk populations[16, 17, 26–28]. Accordingly, simply replicating a CTI-stroke association offers limited novelty. A clinically relevant unresolved question is whether the published CTI formula transports to the U.S. general population and adds measurable information beyond age, socioeconomic factors, obesity, diabetes, hypertension, smoking, TyG, or its individual components.

Using NHANES 1999-2010 data, we assessed the transportability and incremental value of CTI in relation to prevalent stroke among U.S. adults while accounting for the complex survey design and fasting subsample weights. Rather than treating an observed association as validation, we examined whether the published formula retained information beyond established risk factors. We therefore evaluated adjusted associations and dose-response, reported subgroup estimates with formal interaction tests, performed sensitivity analysis excluding CRP >10 mg/L, compared CTI with TyG and its components, and quantified incremental discrimination beyond traditional cardiometabolic covariates.

## Materials and methods

### Study design and population

Data were obtained from NHANES 1999-2010. NHANES is a continuous, nationally representative survey of the civilian noninstitutionalized U.S. population that uses a complex, multistage probability sampling design. NHANES protocols were approved by the National Center for Health Statistics Research Ethics Review Board, and all participants provided written informed consent. The present study used publicly available, de-identified data and was conducted as a cross-sectional analysis of prevalent stroke. The data were accessed for research purposes on 9 June 2026. The authors did not have access to information that could identify individual participants during or after data collection.

We restricted the analysis to 1999-2010 because conventional CRP and the fasting triglyceride and glucose components required for the same CTI definition were jointly available across these six cycles. CRP was not released in the 2011-2014 cycles, and high-sensitivity CRP was reintroduced in 2015-2016 as a new component measured with a different assay framework[29, 30]. Later cycles were therefore not pooled to avoid introducing cross-era CRP measurement heterogeneity.

A total of 62,160 participants from six NHANES cycles were screened. We excluded 29,696 participants aged <20 years, 56 with missing stroke status, 18,447 with missing CRP, triglyceride, or fasting glucose data required for CTI calculation, 1,147 with missing fasting subsample weights, and 1,005 with missing family income-to-poverty ratio. The analytic cohort therefore included 11,809 of 32,464 adults initially screened (36.4%). Most adult exclusions reflected unavailable fasting CTI components or survey weights. Regression models used complete cases for their included covariates. Survey weighting supports population inference for eligible observations but cannot fully correct selection related to laboratory availability or complete-case inclusion. The participant selection process is shown in Figure 1.

**Figure 1.**
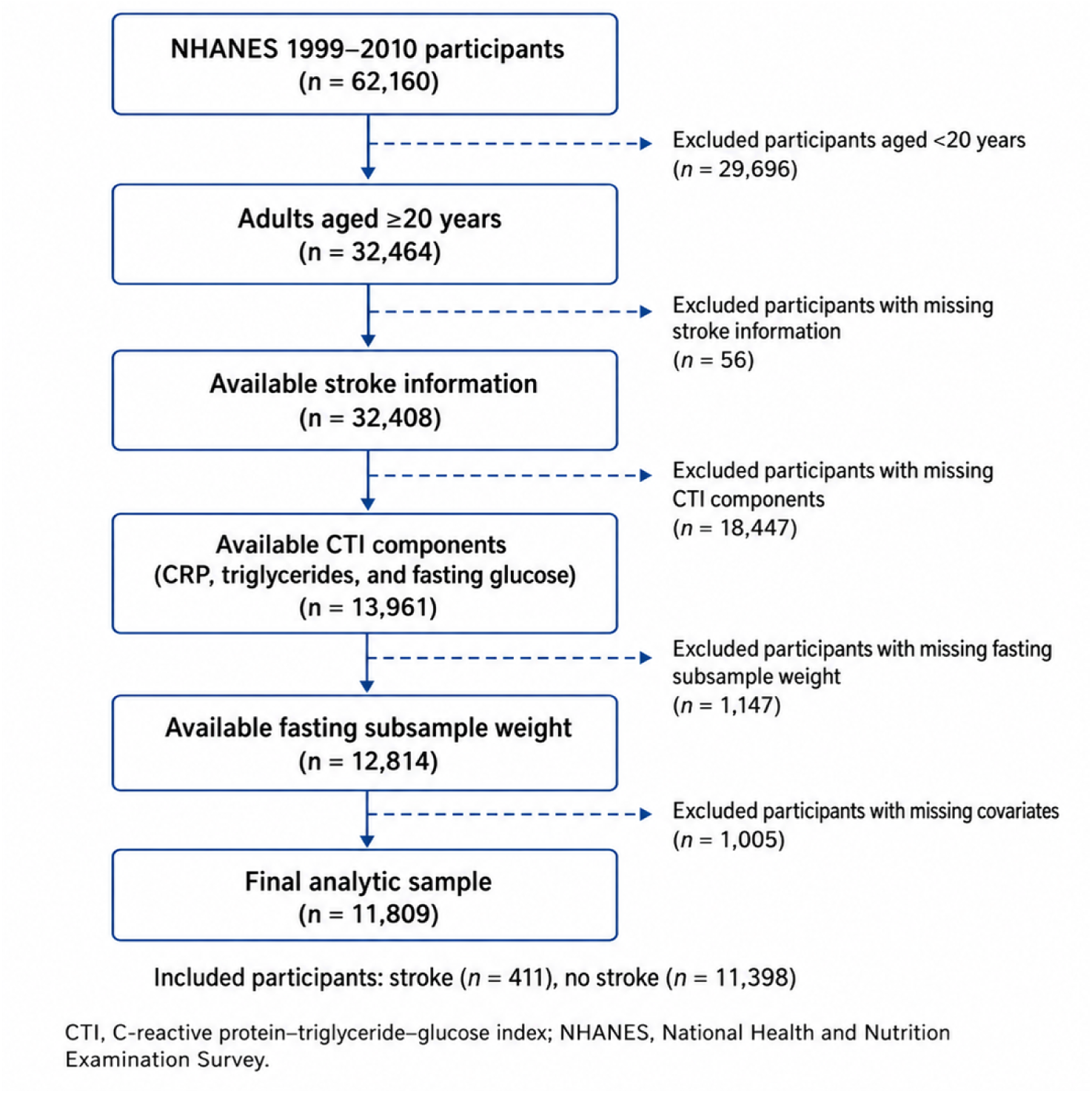
Flow diagram of participant selection from NHANES 1999–2010. Note: A total of 62,160 participants from NHANES 1999-2010 were initially screened. Sequential exclusions were age <20 years (n = 29,696), missing stroke information (n = 56), missing CTI components (n = 18,447), missing fasting subsample weights (n = 1,147), and missing family income-to-poverty ratio (n = 1,005). The final analytic cohort included 11,809 adults, including 411 participants with and 11,398 without a history of stroke. CTI, C-reactive protein-triglyceride-glucose index; NHANES, National Health and Nutrition Examination Survey.

### Assessment of CTI

CTI was calculated using serum CRP, triglycerides, and fasting glucose measured with standardized NHANES laboratory methods. We applied the published formula without re-estimating its coefficients[10]:

CTI = 0.412 × ln(CRP [mg/L]) + ln(TG [mg/dL] × FPG [mg/dL]) / 2 Ruan et al. developed CTI in 5,221 patients with cancer by combining the TyG term with a prognostic weight of 0.412 for ln(CRP)[10]. Thus, 0.412 is an empirically derived coefficient from a cancer-survival model rather than a biological constant. We retained the original coefficient to test the published index as defined, but did not recalibrate it for the U.S. general population or demographic subgroups.

Triglycerides and fasting glucose were measured in the fasting subsample; therefore, fasting subsample weights were used in all weighted analyses. Because NHANES reported CRP in mg/dL during these cycles, CRP values were converted to mg/L before CTI calculation.

### Definition of stroke

Stroke status was determined using the medical conditions questionnaire. Participants were classified as having prevalent stroke if they answered "yes" to the question asking whether a doctor or other health professional had ever told them that they had had a stroke. Participants who answered "no" were classified as not having a history of stroke.

### Covariates

Potential confounders were selected according to previous literature and clinical relevance. Demographic covariates included age, sex, race/ethnicity, education level, and family income-to-poverty ratio. Race/ethnicity was categorized as Mexican American, Other Hispanic, Non-Hispanic White, Non-Hispanic Black, and Other Race. Education level was categorized as less than 9th grade, 9-11th grade, high school graduate/GED, some college or associate degree, and college graduate or above.

Cardiometabolic and lifestyle covariates included body mass index (BMI), diabetes, hypertension, and smoking status. BMI was calculated as weight in kilograms divided by height in meters squared. Diabetes and hypertension were defined according to self-reported physician diagnosis. Smoking status was categorized as never, former, or current smoker according to questionnaire data.

### Statistical analysis

All analyses accounted for the NHANES complex sampling design, including sampling weights, strata, and primary sampling units[19–21]. Because CTI was calculated from fasting triglycerides and fasting glucose, fasting subsample weights were applied. For the combined NHANES 1999-2010 cycles, the 2-year fasting subsample weight (WTSAF2YR) was divided by six to generate the 12-year fasting subsample weight.

Continuous variables are presented as weighted means with standard errors, and categorical variables are presented as numbers with weighted percentages. Baseline characteristics were compared across CTI quartiles, including age and cardiometabolic features. CTI was analyzed as a continuous variable and as a categorical variable according to quartiles. Survey-weighted logistic regression was used to evaluate associations with prevalent stroke, and results are reported as ORs with 95% CIs.

Three regression models were constructed to separate demographic adjustment from adjustment for cardiometabolic and lifestyle factors. Model 1 was unadjusted. Model 2 was adjusted for age, sex, race/ethnicity, education level, and family income-to-poverty ratio. Model 3 was further adjusted for BMI, diabetes, hypertension, and smoking status. It included 11,344 complete cases, excluding 465 participants (3.9% of the analytic cohort) with at least one additional Model 3 covariate missing. Model 3 addressed whether CTI retained information beyond established risk factors. It was not interpreted as an etiologic total-effect model because diabetes and hypertension may lie on pathways linking inflammatory-metabolic burden with stroke.

Exploratory subgroup analyses were performed using age strata of <60 and > =60 years and strata defined by sex, BMI (< 30 or > =30 kg/m^2^), diabetes status, and hypertension status. Multiplicative interaction terms between CTI and each subgroup variable were added to the fully adjusted model. Evidence of effect modification was evaluated from the interaction term rather than from differences in statistical significance between subgroup-specific estimates.

Restricted cubic spline analysis was performed to explore the shape of the CTI-stroke association. Four knots were placed at the 5th, 35th, 65th, and 95th percentiles of the CTI distribution, and the median CTI value was used as the reference. The spline model was fitted using survey-weighted logistic regression and adjusted for age, sex, race/ethnicity, education level, family income-to-poverty ratio, BMI, diabetes, hypertension, and smoking status. The overall association and non-linearity were evaluated using Wald tests.

To examine comparative and incremental value, we fitted a traditional covariate model with age, sex, race/ethnicity, education level, family income-to-poverty ratio, BMI, diabetes, hypertension, and smoking status. We then separately added CTI, TyG, ln(CRP), ln(triglycerides), and ln(fasting glucose). Biomarkers were standardized, and ORs are reported per 1-SD increase. Incremental discrimination was assessed using weighted C-statistics, change in C-statistic, IDI, and continuous NRI[22]. IDI and NRI were interpreted together with the change in C-statistic and were not treated as independent evidence of clinically meaningful improvement. Because acute inflammatory states can elevate CRP, sensitivity analysis repeated the fully adjusted CTI models after restricting to participants with CRP <=10 mg/L. We also compared key baseline characteristics of participants at or below versus above this threshold using survey-weighted tests (Supplementary Table S1).

All statistical analyses were performed using R software (version 4.3.1). A two-sided P value <0.05 was considered statistically significant.

## Results

### Study population

A total of 62,160 participants from NHANES 1999-2010 were initially screened. Sequential exclusions were age <20 years (n = 29,696), missing stroke information (n = 56), missing CTI components (n = 18,447), missing fasting subsample weights (n = 1,147), and missing family income-to-poverty ratio (n = 1,005). The final analytic cohort included 11,809 adults, which was 36.4% of the 32,464 adults initially screened. Among them, 411 reported a history of stroke and 11,398 reported no history of stroke (Figure 1). The fully adjusted Model 3 included 11,344 participants, or 96.1% of the analytic cohort.

### Baseline characteristics

The 11,809 participants were categorized into four CTI quartiles: Q1 (CTI ≤ 4.565), Q2 (4.565 < CTI ≤ 5.042), Q3 (5.042 < CTI ≤ 5.493), and Q4 (CTI > 5.493). Baseline characteristics across CTI quartiles are presented in Table 1.

**Table 1.** Baseline Characteristics by CTI Quartile Note: Values are presented as weighted means with standard errors for continuous variables and numbers with weighted percentages for categorical variables, unless otherwise specified. CTI quartiles were defined as Q1 (CTI ≤ 4.565), Q2 (4.565 < CTI ≤ 5.042), Q3 (5.042 < CTI ≤ 5.493), and Q4 (CTI > 5.493). CTI was calculated as 0.412 × ln(CRP [mg/L]) + ln(TG [mg/dL] × FPG [mg/dL]) / 2. Because CRP values in NHANES are reported in mg/dL, they were converted to mg/L before CTI calculation. P values were calculated using survey-weighted tests. CTI, C-reactive protein–triglyceride–glucose index; CRP, C-reactive protein; TG, triglycerides; FPG, fasting plasma glucose; PIR, family income-to-poverty ratio; BMI, body mass index; HDL-C, high-density lipoprotein cholesterol; TC, total cholesterol; HbA1c, glycated hemoglobin; NHANES, National Health and Nutrition Examination Survey.

| Var. | Group | Q1 | Q2 | Q3 | Q4 | P |
| --- | --- | --- | --- | --- | --- | --- |
| N |  | 2953 | 2952 | 2952 | 2952 |  |
| Age, y |  | 40.66 (15.42) | 46.71 (16.52) | 48.16 (16.92) | 49.45 (16.13) | <0.001 |
| Sex | Male | 26844749.8 (48.5) | 26917235.4 (54.4) | 21905114.8 (48.8) | 17000154.4 (40.7) | <0.001 |
|  | Female | 28513537.1 (51.5) | 22524531.4 (45.6) | 22969259.4 (51.2) | 24783823.8 (59.3) |  |
| Race/ethn. | Mexican American | 3389188.1 (6.1) | 3451682.3 (7.0) | 3987422.7 (8.9) | 3623975.2 (8.7) | <0.001 |
|  | Other Hispanic | 2124636.9 (3.8) | 2565075.0 (5.2) | 2339424.6 (5.2) | 1957928.7 (4.7) |  |
|  | Non-Hispanic White | 39387710.8 (71.2) | 35717938.2 (72.2) | 31868750.3 (71.0) | 30156717.4 (72.2) |  |
|  | Non-Hispanic Black | 6473025.3 (11.7) | 5164295.8 (10.4) | 4418714.4 (9.8) | 4338360.5 (10.4) |  |
|  | Other Race | 3983725.8 (7.2) | 2542775.6 (5.1) | 2260062.2 (5.0) | 1706996.3 (4.1) |  |
| Education | Less than 9th grade | 2243060.0 (4.1) | 3224321.3 (6.5) | 3628247.0 (8.1) | 3373448.1 (8.1) | <0.001 |
|  | 9–11th grade | 5257095.4 (9.5) | 5915241.8 (12.0) | 5584251.6 (12.5) | 6712338.2 (16.1) |  |
|  | High school/GED | 11731583.2 (21.2) | 12594041.9 (25.5) | 11929830.8 (26.6) | 11620991.6 (27.9) |  |
|  | Some college/AA | 16988767.9 (30.7) | 14612679.3 (29.6) | 13804695.9 (30.8) | 12578115.4 (30.2) |  |
|  | College graduate or above | 19109571.4 (34.5) | 13053504.1 (26.4) | 9897910.0 (22.1) | 7417462.9 (17.8) |  |
| PIR |  | 3.21 (1.60) | 3.10 (1.59) | 2.96 (1.60) | 2.77 (1.63) | <0.001 |
| BMI, kg/m<sup>2</sup> |  | 24.56 (4.16) | 27.56 (5.03) | 29.91 (6.11) | 32.95 (7.58) | <0.001 |
| Diabetes | No | 53511723.0 (97.4) | 46350428.9 (94.8) | 40513196.3 (91.9) | 34786315.8 (85.0) | <0.001 |
|  | Yes | 1424651.2 (2.6) | 2564221.6 (5.2) | 3579949.3 (8.1) | 6155596.3 (15.0) |  |
| Hypertension | No | 46655436.9 (84.6) | 34997075.5 (71.2) | 29808384.6 (66.7) | 23441508.1 (56.3) | <0.001 |
|  | Yes | 8486490.8 (15.4) | 14144634.4 (28.8) | 14914706.3 (33.3) | 18198221.0 (43.7) |  |
| Smoking | Never | 31963847.1 (57.7) | 25854005.9 (52.3) | 22230742.4 (49.6) | 18640475.0 (44.7) | <0.001 |
|  | Former | 12150077.7 (22.0) | 12356332.9 (25.0) | 11911337.9 (26.6) | 12329267.3 (29.5) |  |
|  | Current | 11238557.9 (20.3) | 11231428.0 (22.7) | 10689700.5 (23.8) | 10767660.9 (25.8) |  |
| Stroke | No stroke | 54727098.2 (98.9) | 48245491.7 (97.6) | 43642816.4 (97.3) | 39905989.6 (95.5) | <0.001 |
|  | Stroke | 631188.7 (1.1) | 1196275.1 (2.4) | 1231557.7 (2.7) | 1877988.5 (4.5) |  |
| CRP, mg/L |  | 0.60 (0.43) | 1.81 (1.14) | 3.92 (2.63) | 12.19 (14.41) | <0.001 |
| TG, mg/dL |  | 82.49 (35.60) | 118.57 (53.99) | 155.31 (89.47) | 227.17 (210.71) | <0.001 |
| FPG,<br>mg/dL |  | 94.62 (13.24) | 98.92 (15.06) | 103.16 (23.18) | 116.24 (46.10) | <0.001 |
| HDL-C,<br>mg/dL |  | 59.83 (15.86) | 53.36 (14.96) | 50.15 (14.60) | 46.94 (14.09) | <0.001 |
| TC, mg/dL |  | 185.95 (35.60) | 196.44 (38.25) | 204.59 (40.41) | 211.73 (48.61) | <0.001 |
| HbA1c, % |  | 5.24 (0.48) | 5.38 (0.53) | 5.52 (0.74) | 5.92 (1.34) | <0.001 |
| Uric acid,<br>mg/dL |  | 4.95 (1.25) | 5.49 (1.34) | 5.68 (1.39) | 5.88 (1.51) | <0.001 |
| Creatinine,<br>mg/dL |  | 0.85 (0.20) | 0.88 (0.39) | 0.87 (0.33) | 0.88 (0.52) | <0.001 |

**Note:** Values are presented as weighted means with standard errors for continuous variables and numbers with weighted percentages for categorical variables, unless otherwise specified. CTI quartiles were defined as Q1 ( $CTI \leq 4.565$ ), Q2 ( $4.565 < CTI \leq 5.042$ ), Q3 ( $5.042 < CTI$ $\leq 5.493$ ), and Q4 ( $CTI > 5.493$ ). CTI was calculated as $0.412 \times \ln(CRP$ $[mg/L]) + \ln(TG [mg/dL] \times FPG [mg/dL]) / 2$ . Because CRP values in NHANES are reported in mg/dL, they were converted to mg/L before CTI calculation. P values were calculated using survey-weighted tests. CTI, C-reactive protein–triglyceride–glucose index; CRP, C-reactive protein; TG, triglycerides; FPG, fasting plasma glucose; PIR, family income-to-poverty ratio; BMI, body mass index; HDL-C, high-density lipoprotein cholesterol;

Higher CTI quartiles were characterized by a progressively less favorable inflammatory-metabolic profile. Participants in higher quartiles tended to be older and had higher BMI values. The weighted prevalence of diabetes, hypertension, and stroke increased across CTI quartiles, and socioeconomic and smoking distributions also differed across groups.

As expected from the CTI formula, CRP, triglycerides, and fasting glucose increased across quartiles. Other metabolic and vascular markers also showed adverse patterns. HbA1c, uric acid, serum creatinine, and total cholesterol were generally higher, whereas HDL cholesterol was lower in higher CTI quartiles. These gradients support interpreting CTI as a compact marker of systemic inflammatory-metabolic burden rather than a single-pathway biomarker.

### Association between CTI and prevalent stroke

The association between CTI and prevalent stroke is presented in Table 2. When CTI was analyzed as a continuous variable, higher CTI was associated with stroke in the unadjusted model (Model 1: OR = 2.05, 95% CI: 1.74-2.42, P < 0.001). The association remained statistically significant after adjustment for age, sex, race/ethnicity, education level, and family income-to-poverty ratio (Model 2: OR = 1.57, 95% CI: 1.31-1.89, P < 0.001). After further adjustment for BMI, diabetes, hypertension, and smoking status, the estimate was attenuated and no longer statistically significant (Model 3: OR = 1.22, 95% CI: 0.98-1.53, P = 0.080).

**Table 2.**
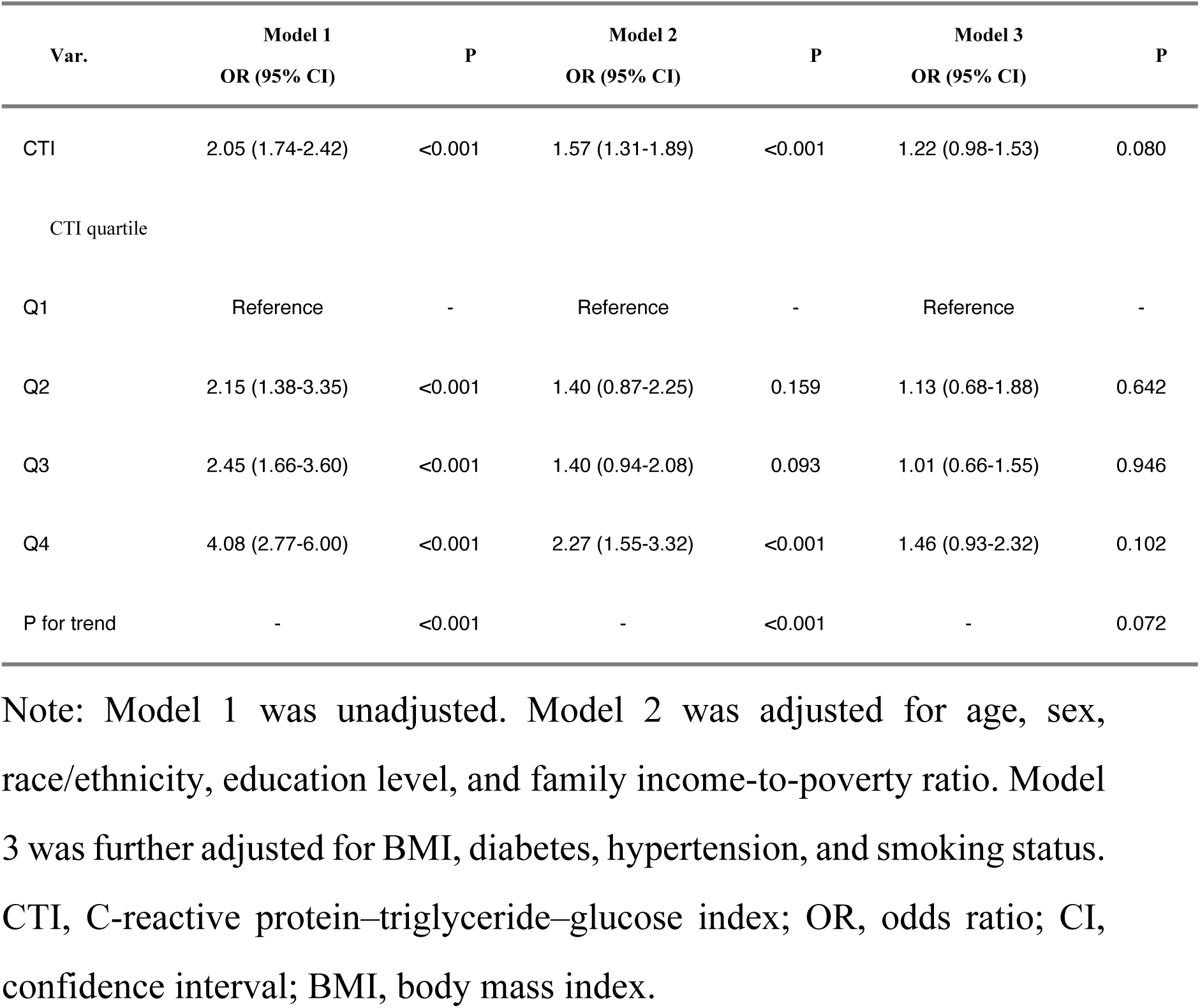
Association Between CTI and Prevalent Stroke Note: Model 1 was unadjusted. Model 2 was adjusted for age, sex, race/ethnicity, education level, and family income-to-poverty ratio. Model 3 was further adjusted for BMI, diabetes, hypertension, and smoking status. CTI, C-reactive protein–triglyceride–glucose index; OR, odds ratio; CI, confidence interval; BMI, body mass index.

Quartile-based analyses showed a similar pattern. In Model 1, compared with Q1, the ORs were 2.15 (95% CI: 1.38-3.35) for Q2, 2.45 (95% CI: 1.66-3.60) for Q3, and 4.08 (95% CI: 2.77-6.00) for Q4. In Model 2, the association remained significant for Q4 (OR = 2.27, 95% CI: 1.55-3.32, P < 0.001). In Model 3, however, the quartile associations were not statistically significant: OR = 1.13 (95% CI: 0.68-1.88) for Q2, 1.01 (95% CI: 0.66-1.55) for Q3, and 1.46 (95% CI: 0.93-2.32) for Q4. The trend across CTI quartiles did not reach statistical significance after full adjustment (P for trend = 0.072).

### Exploratory subgroup and interaction analyses

Exploratory subgroup analyses are shown in Table 3 and Figure 2. In the fully adjusted models, the CTI estimate was OR = 1.30 (95% CI: 1.01-1.66; nominal P = 0.045) among participants aged >=60 years and OR = 1.24 (95% CI: 0.81-1.90; P = 0.323) among those aged <60 years. However, the CTI-by-age interaction was not statistically significant (P interaction = 0.287); therefore, the difference between subgroup-specific P values does not constitute evidence of age-related effect modification. The estimate among participants without diabetes was OR = 1.40 (95% CI: 1.04-1.88; nominal P = 0.028), but the diabetes interaction and all other interaction tests were also non-significant. These subgroup observations are hypothesis-generating only.

**Figure 2.**
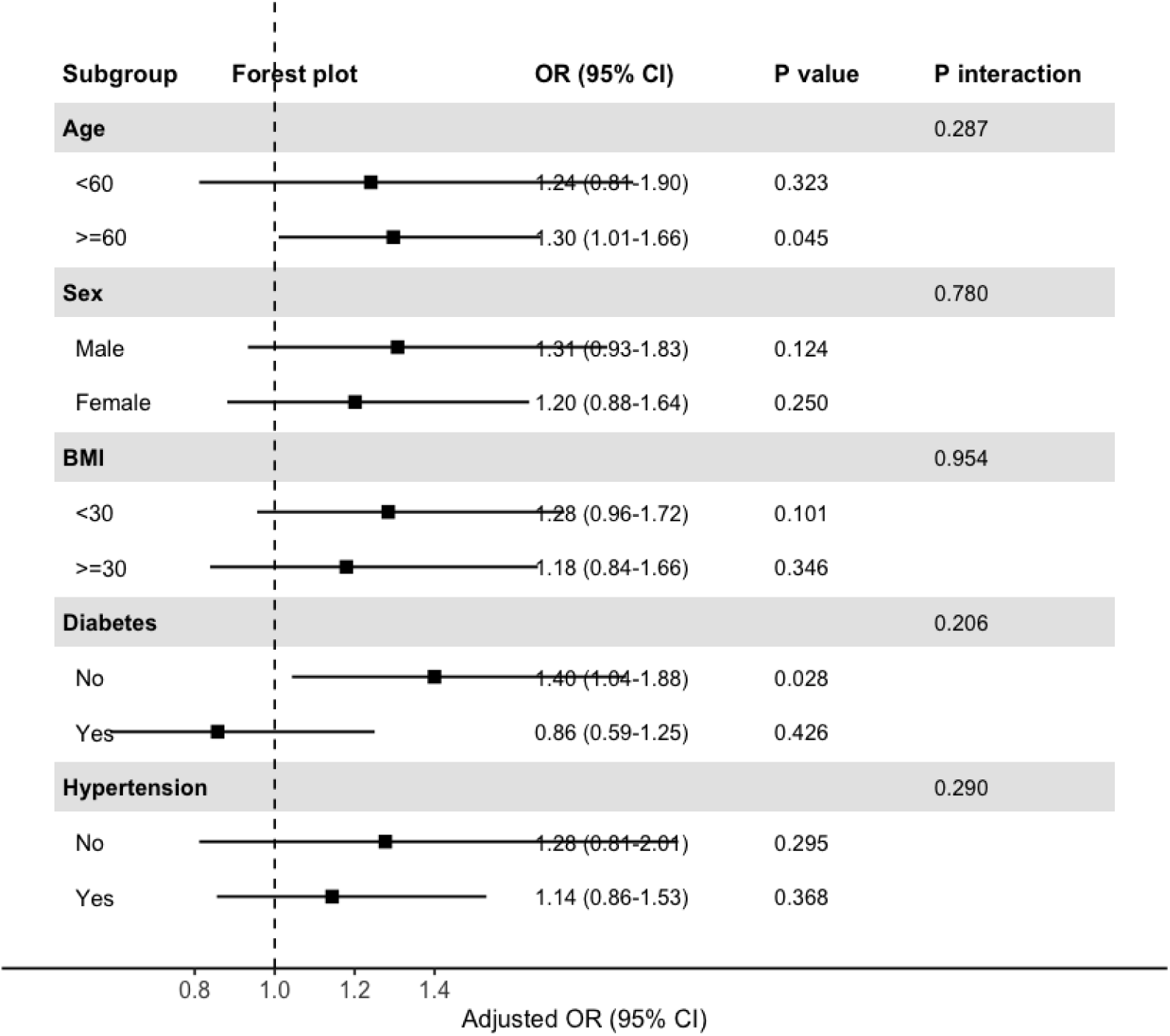
Exploratory subgroup analyses of the association between CTI and prevalent stroke. Note: The forest plot shows the adjusted odds ratios and 95% confidence intervals for the association between CTI and prevalent stroke across subgroups defined by age, sex, BMI, diabetes status, and hypertension status. Square points represent odds ratios, and horizontal lines represent 95% confidence intervals. The vertical dashed line indicates an odds ratio of 1. Models were adjusted for age, sex, race/ethnicity, education level, family income-to-poverty ratio, BMI, diabetes, hypertension, and smoking status, except for the corresponding stratification variable. CTI, C-reactive protein–triglyceride–glucose index; BMI, body mass index; OR, odds ratio; CI, confidence interval.

**Table 3.** Exploratory subgroup analyses of CTI and prevalent stroke **Note:** Odds ratios and 95% confidence intervals were estimated using fully adjusted survey-weighted logistic regression models. Models were adjusted for age, sex, race/ethnicity, education level, family income-to-poverty ratio, BMI, diabetes, hypertension, and smoking status, except for the corresponding stratification variable. CTI, C-reactive protein–triglyceride–glucose index; OR, odds ratio; CI, confidence interval; BMI, body mass index.

| Subgroup | OR (95% CI) | P | P<br>interaction |
| --- | --- | --- | --- |
| Age |  |  | 0.287 |
| <60 years | 1.24 (0.81-1.90) | 0.323 |  |
| ≥60 years | 1.30 (1.01-1.66) | 0.045 |  |
| Sex |  |  | 0.780 |
| Male | 1.31 (0.93-1.83) | 0.124 |  |
| Female | 1.20 (0.88-1.64) | 0.250 |  |
| BMI |  |  | 0.954 |
| <30 kg/m<sup>2</sup> | 1.28 (0.96-1.72) | 0.101 |  |
| ≥30 kg/m<sup>2</sup> | 1.18 (0.84-1.66) | 0.346 |  |
| Diabetes |  |  | 0.206 |
| No | 1.40 (1.04-1.88) | 0.028 |  |
| Yes | 0.86 (0.59-1.25) | 0.426 |  |
| Hypertension |  |  | 0.290 |
| No | 1.28 (0.81-2.01) | 0.295 |  |
| Yes | 1.14 (0.86-1.53) | 0.368 |  |

### Restricted cubic spline analysis

Restricted cubic spline analysis was used to evaluate the dose-response pattern between CTI and prevalent stroke (Figure 3). The spline curve suggested a generally increasing trend in the odds of stroke at higher CTI levels. However, the overall association was not statistically significant (P for overall association = 0.369), and there was no evidence of non-linearity (P for non-linearity = 0.996). These findings are consistent with the fully adjusted regression models and do not support a strong threshold or non-linear association.

**Figure 3.**
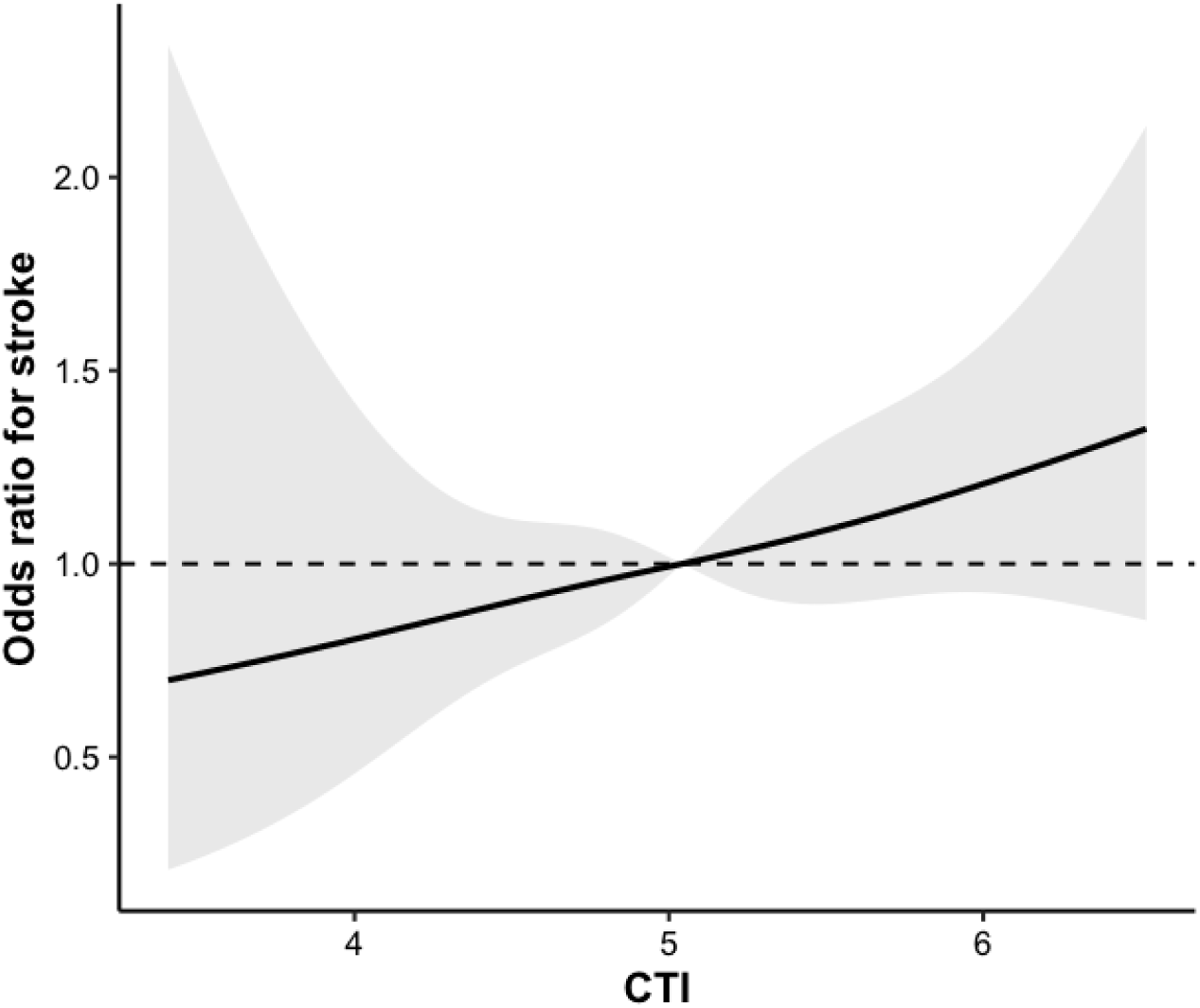
Restricted cubic spline analysis of the association between CTI and prevalent stroke. Note: Restricted cubic spline analysis was performed to evaluate the dose-response relationship between CTI and prevalent stroke. The solid line represents the adjusted odds ratio for stroke according to CTI levels, and the shaded area represents the 95% confidence interval. The horizontal dashed line indicates an odds ratio of 1. The median CTI value was used as the reference. The model was adjusted for age, sex, race/ethnicity, education level, family income-to-poverty ratio, BMI, diabetes, hypertension, and smoking status. The overall association was not statistically significant (P for overall association = 0.369), and no significant non-linear association was observed (P for non-linearity = 0.996). CTI, C-reactive protein–triglyceride–glucose index; OR, odds ratio; CI, confidence interval.

### Comparative biomarker value and acute-inflammation sensitivity

In the complete-case sample used for fully adjusted model comparison (n = 11,344; 374 stroke events), the traditional covariate model had a weighted C-statistic of 0.846 (Table 4). Adding CTI increased the C-statistic to 0.847 (Delta C = 0.001), with an IDI of 0.0003 and a continuous NRI of 0.101. The NRI was not accompanied by a meaningful change in overall discrimination or IDI and was not interpreted as evidence of clinical improvement. CTI therefore provided negligible incremental discrimination beyond age, socioeconomic factors, BMI, diabetes, hypertension, smoking, and other established covariates. TyG, ln(triglycerides), and ln(fasting glucose) were not statistically associated with prevalent stroke after full adjustment. ln(CRP) was associated with prevalent stroke (OR per 1 SD = 1.17, 95% CI: 1.02-1.34, P = 0.032), but its addition also increased the C-statistic only to 0.847.

**Table 4.**
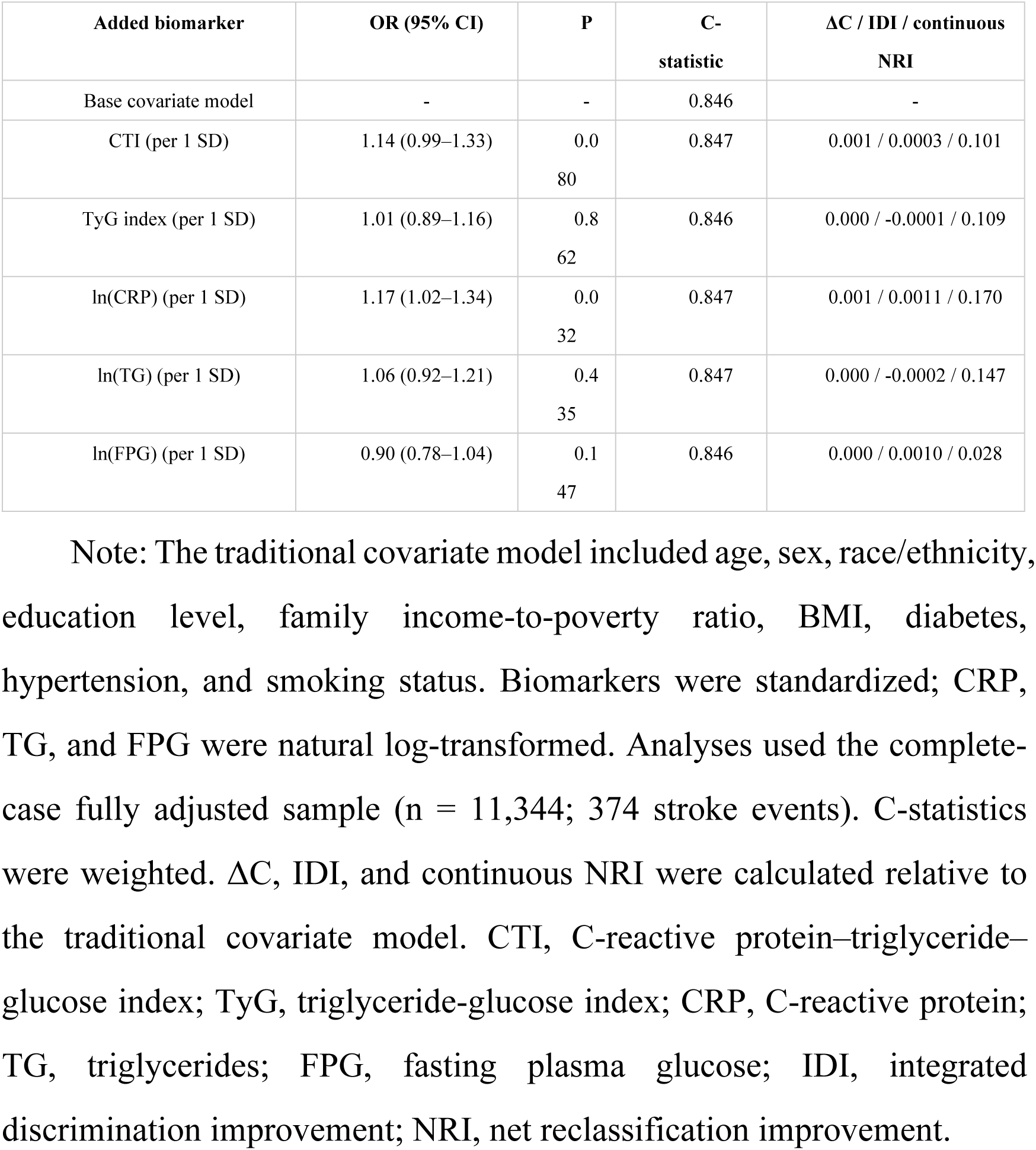
Comparative and incremental value of CTI, TyG, and component biomarkers for prevalent stroke Note: The traditional covariate model included age, sex, race/ethnicity, education level, family income-to-poverty ratio, BMI, diabetes, hypertension, and smoking status. Biomarkers were standardized; CRP, TG, and FPG were natural log-transformed. Analyses used the complete-case fully adjusted sample (n = 11,344; 374 stroke events). C-statistics were weighted. ΔC, IDI, and continuous NRI were calculated relative to the traditional covariate model. CTI, C-reactive protein–triglyceride–glucose index; TyG, triglyceride-glucose index; CRP, C-reactive protein; TG, triglycerides; FPG, fasting plasma glucose; IDI, integrated discrimination improvement; NRI, net reclassification improvement.

Among the 11,809 analytic participants, 10,457 had CRP <=10 mg/L and 1,352 had CRP >10 mg/L. Compared with participants at or below the threshold, those above it were older, more often women, had higher BMI, and had higher weighted prevalences of diabetes, hypertension, current smoking, and prevalent stroke (4.2% vs. 2.4%; Supplementary Table S1). After the CRP restriction and complete-case covariate filtering, the fully adjusted sensitivity model included 10,063 participants and 313 stroke events (Table 5). The association remained non-significant when CTI was modeled continuously (OR = 1.18, 95% CI: 0.88-1.58, P = 0.264), per 1-SD increase (OR = 1.11, 95% CI: 0.93-1.32, P = 0.264), and by quartiles (P for trend = 0.321). This analysis tests robustness to extreme CRP values but cannot eliminate selection bias because the excluded and retained groups differed systematically.

**Table 5.** Sensitivity analysis excluding participants with CRP >10 mg/L Note: After restricting the analytic cohort to CRP <=10 mg/L and applying complete-case requirements for the fully adjusted model, the sensitivity sample included 10,063 participants and 313 stroke events. Quartiles were recalculated within this sensitivity-model sample. Models were adjusted for age, sex, race/ethnicity, education level, family income-to-poverty ratio, BMI, diabetes, hypertension, and smoking status. CRP, C-reactive protein; CTI, C-reactive protein-triglyceride-glucose index; OR, odds ratio; CI, confidence interval. Supplementary Table S1. Characteristics of participants according to the CRP sensitivity-analysis threshold Note: The threshold comparison includes 10,457 participants with CRP <=10 mg/L and 1,352 participants with CRP >10 mg/L. Values are weighted means (SE) for continuous variables and unweighted n (survey-weighted %) for categorical variables. The table is provided as a separate Excel file.

| Biomarker | OR (95% CI) | P |
| --- | --- | --- |
| CTI (per 1-unit increase) | 1.18 (0.88–1.58) | 0.264 |
| CTI (per 1 SD) | 1.11 (0.93–1.32) | 0.264 |
| Q1 | Reference | - |
| Q2 | 1.30 (0.75–2.26) | 0.357 |
| Q3 | 0.97 (0.59–1.61) | 0.920 |
| Q4 | 1.40 (0.83–2.36) | 0.211 |
| P for trend | - | 0.321 |

Note: After restricting the analytic cohort to CRP ≤10 mg/L and applying complete-case requirements for the fully adjusted model, the sensitivity sample included 10,063 participants and 313 stroke events. Quartiles were recalculated within this sensitivity-model sample. Models were adjusted for age, sex, race/ethnicity, education level, family income-to-poverty ratio, BMI, diabetes, hypertension, and smoking status. CRP, C-reactive protein; CTI, C-reactive protein-triglyceride-glucose index; OR, odds ratio; CI, confidence interval.

## Discussion

In this survey-weighted NHANES 1999-2010 analysis, CTI identified adults with a higher inflammatory-metabolic burden and was associated with prevalent stroke before full cardiometabolic adjustment. The fully adjusted association was not statistically significant, all formal subgroup interaction tests were non-significant, and CTI did not materially improve discrimination beyond traditional covariates. The study therefore assesses the transportability and incremental value of the published formula rather than providing comprehensive external or clinical validation.

The following pathways provide biological plausibility for the observed association, but the cross-sectional design precludes causal inference. CTI combines CRP, triglycerides, and fasting glucose[10], which index systemic inflammation, lipid metabolism, and glucose metabolism. These domains have been implicated in atherosclerosis, endothelial dysfunction, insulin resistance, thrombotic activation, and vascular injury[23–25]. These abnormalities often co-occur and may reinforce one another. However, the gradients across CTI quartiles show correlation only and do not establish that CTI or its components caused stroke.

Our findings should be interpreted alongside recent studies of CTI and stroke or cardiovascular outcomes. Prospective CHARLS analyses have reported associations of baseline CTI, cumulative exposure, and longitudinal trajectories with incident stroke, including variation by glycemic status[17, 26]. Other prospective work has related CTI to cardiovascular events or mortality across cardiovascular-kidney-metabolic stages[27], and NHANES studies have examined CTI in selected coronary heart disease and diabetes populations[28]. Our analysis does not establish antecedent risk because CTI and stroke history were measured concurrently. It instead assesses whether the published score retains information beyond traditional covariates in a general U.S. sample. The negligible changes in C-statistic and IDI argue against meaningful added discrimination.

The incremental analyses support the main interpretation of this study. A composite biomarker can reflect inflammatory-metabolic burden yet remain clinically redundant when it overlaps with age, obesity, diabetes, hypertension, smoking, and socioeconomic factors. The traditional covariate model already showed high discrimination for prevalent stroke. Adding CTI increased the weighted C-statistic by only 0.001, and the IDI was close to zero. Although continuous NRI was 0.101, it was not accompanied by meaningful improvement in overall discrimination and can be sensitive to small probability changes[22]. Adjustment for diabetes and hypertension may remove part of an etiologic pathway, so Model 3 should not be read as a total causal-effect estimate. Their inclusion remains appropriate for the separate question of incremental information beyond established risk factors.

Subgroup findings require careful interpretation. Nominal associations were observed among adults aged >=60 years and participants without diabetes, but neither the age nor diabetes interaction term was statistically significant. Differences in whether subgroup-specific P values cross 0.05 do not establish differences between groups. These results provide no evidence of age-specific or diabetes-specific effects and should be considered exploratory.

The spline analysis also supports a cautious interpretation. Although the curve trended upward, neither the overall association nor the non-linear component was statistically significant after full adjustment. The findings therefore do not support a clear CTI threshold for stroke prevalence. Prospective studies should examine repeated CTI measurements, incident stroke, and direct vascular measures before drawing conclusions about vascular aging or clinical risk.

This study has several strengths. First, NHANES provides a nationally representative sample of the U.S. civilian noninstitutionalized population. Second, analyses accounted for the complex survey design and fasting subsample weights. Third, CTI was evaluated continuously, by quartiles, and per 1-SD increase. Fourth, subgroup estimates were paired with formal interaction tests to avoid interpreting isolated subgroup P values as effect modification. Fifth, CTI was benchmarked against TyG and its individual components and evaluated for incremental discrimination beyond traditional cardiometabolic covariates. Sixth, the acute-inflammation sensitivity analysis was accompanied by a comparison of participants above and below the CRP threshold.

Several limitations should be acknowledged. First, the cross-sectional design and prevalent-stroke outcome preclude temporal or causal inference. Stroke survivors may experience long-term changes in inflammation, glucose and lipid metabolism, medication use, diet, and physical activity; consequently, elevated CTI may be a consequence of stroke or post-stroke care rather than an antecedent risk marker. Second, stroke status was based on self-reported physician diagnosis and may be affected by recall error or misclassification. Third, CTI was calculated from single biomarker measurements, which may not represent long-term inflammatory-metabolic burden.

Fourth, the coefficient 0.412 was derived in a cancer-survival cohort and was not recalibrated in NHANES. We also did not assess calibration, so this study evaluates transportability and incremental value rather than comprehensive external validation. Fifth, adjustment for diabetes and hypertension may partly overadjust an etiologic association, although it is appropriate for evaluating incremental information. Sixth, the complete-case design and large exclusions for unavailable fasting biomarkers may limit representativeness. Survey weights cannot eliminate selection bias caused by missingness. The CRP >10 mg/L restriction tested robustness to extreme values, but systematic differences between excluded and retained participants mean that it also cannot eliminate selection bias. Seventh, NHANES 1999-2010 used conventional CRP rather than contemporary hs-CRP. Differences in analytic sensitivity and measurement range may limit the transportability of this CTI formula to current clinical practice. Residual confounding from medication use, stroke subtype, disease duration, diet, physical activity, infection, chronic inflammatory disease, frailty, renal function, and other factors also remains possible. Finally, all subgroup findings were exploratory because interaction tests were non-significant.

Taken together, CTI reflected correlated inflammatory-metabolic burden and was associated with prevalent stroke before full cardiometabolic adjustment. The cross-sectional design cannot distinguish antecedent risk from post-stroke metabolic alterations. Attenuation after adjustment, non-significant interaction tests, and negligible incremental discrimination do not support CTI as a clinically useful stand-alone stroke marker in NHANES.

## Conclusion

In this survey-weighted NHANES 1999-2010 analysis, CTI reflected correlated inflammatory-metabolic burden, but its association with prevalent stroke was largely accounted for by traditional risk factors. The study evaluates transportability and incremental value rather than comprehensive external validation. CTI did not materially improve discrimination, and the data do not support its use as a stand-alone clinical stroke marker.

## Data Availability

The datasets analyzed in this study are publicly available from the National Health and Nutrition Examination Survey website.

https://wwwn.cdc.gov/nchs/nhanes/default.aspx

## Declarations

### Ethics approval and consent to participate

The NHANES study protocols were reviewed and approved by the National Center for Health Statistics Research Ethics Review Board. All participants provided written informed consent. The present study used publicly available and de-identified NHANES data and was conducted in accordance with the Declaration of Helsinki. Additional institutional review board approval was not required for this secondary analysis of publicly available de-identified data.

## Consent to participate

All NHANES participants provided written informed consent.

## Clinical trial number

Not applicable.

## Consent for publication

Not applicable.

## Author Contributions

DQL and YZ designed the study and performed the statistical analyses. DQL drafted the initial manuscript. YZ prepared the visualizations and revised the manuscript. All authors read and approved the final manuscript.

